# Structural Inequality in Clean Fuel Access and Acute Respiratory Infection Among Nigerian Children: An Intersectional Multilevel Analysis

**DOI:** 10.64898/2026.03.02.26347442

**Authors:** Kamaldeen Sunkanmi Abdulraheem, Morufat Titilope Omotayo, Blossom Adaeze Maduafokwa, Abdullateef Temitope Abdulazeez, Ibraheem Shola Abdulraheem

## Abstract

**Background:** Acute respiratory infection (ARI) remains a leading cause of morbidity and mortality among children under five in Nigeria. Although polluting cooking fuels are widely considered a key risk factor, their effects may be shaped by broader socioeconomic and geographic conditions. This study examined both individual and structural determinants of ARI and assessed how these factors intersect to pattern risk.

**Methods:** We analysed data from 28,728 children under five in the 2024 Nigeria Demographic and Health Survey. Three ARI definitions were applied. Survey-weighted quasibinomial logistic regression estimated associations between ARI and cooking fuel type, child age and sex, household wealth quintile, residence type, geopolitical zone, and parental education. To examine intersectional patterning, we conducted a Multilevel Analysis of Individual Heterogeneity and Discriminatory Accuracy (MAIHDA), constructing strata defined by combinations of cooking fuel, wealth, residence, and geopolitical zone. The intraclass correlation coefficient (ICC) quantified between-strata variance.

**Results:** Strict ARI prevalence was 1.9%, and 8.3% of children had broader respiratory symptoms. In unadjusted analyses, polluting fuel use was associated with higher odds of respiratory symptoms (OR 1.85, 95% CI 1.43–2.39). After adjustment, this association was substantially attenuated, indicating confounding by structural factors. Child age was the most consistent predictor: children aged 24–59 months had about half the odds of strict ARI compared with infants (aOR 0.53, 95% CI 0.41–0.68). Geopolitical zone showed the strongest overall association. MAIHDA revealed that 9% of total ARI variance lay between intersectional strata (ICC = 0.09), and this variance was not explained by child age or sex. The population-attributable fraction for polluting fuel declined from 41.4% to 12.4% after adjustment.

**Conclusions:** ARI risk among Nigerian children is shaped more by structural and geographic inequalities than by household fuel use alone. Equity-focused, subnational policies addressing intersecting socioeconomic and regional disadvantage are needed to reduce the ARI burden.

## Background

Pneumonia claimed 2.5 million lives worldwide in 2023, representing a 19% increase compared to 2021, with children under five among the most vulnerable populations, accounting for nearly a quarter of all deaths.[1] Sub-Saharan Africa (SSA) experienced a much lower decline in childhood pneumonia deaths than other regions between 2010 and 2023, and now accounts for more than half of all global under-five pneumonia deaths.[2] Household air pollution is a major modifiable risk factor: around a third of the world’s population relies on solid fuels for cooking and heating, almost doubling children’s risk of developing ARI.[3] Despite this, robust global evidence linking household air pollution to poor child health outcomes has not been matched by comparable evidence from sub-Saharan Africa, where the burden of solid fuel use and child mortality is disproportionately high.[4] Nigeria sits at the centre of this burden. Nigeria has the highest absolute numbers of child deaths worldwide and contributes the largest percentage of pneumonia cases and deaths globally, the majority of which are among children under five.[5,6] Despite advancements in ARI management in Nigeria, the country has not yet met global control targets, and the prevalence of ARIs in sub-Saharans countries ranges from 1.9% to 60.2%, with socioeconomic conditions significantly increasing the risk. [7]Though previous studies have identified factors associated with childhood ARI symptoms in Nigeria, progress made in reducing this major burden has remained limited, with associated factors remaining largely unchanged over a decade of DHS data. [8]

Existing literature has largely examined the determinants of ARI in isolation — treating cooking fuel, poverty, geography, and parental education as independent predictors. This approach overlooks the ways in which these factors intersect and accumulate to produce concentrated disadvantage in specific population subgroups. Intersectional approaches are rapidly becoming a popular framework in the study of population health, recognising that health outcomes cannot be adequately understood by considering social positions independently.[9] However, to date, no study in Nigeria has applied intersectional methods to examine how combinations of household energy poverty, wealth, geography, and residence type jointly shape ARI risk in children under five.

Methodologically, standard regression approaches are ill-suited to intersectional analysis because they require pre-specification of all interaction terms and struggle with the sparse data that arises when multiple social categories are combined. Intersectional Multilevel Analysis of Individual Heterogeneity and Discriminatory Accuracy (I-MAIHDA) is an innovative approach for investigating inequalities in health and other outcomes, with conceptual and methodological advantages over conventional single-level regression analysis, enabling the study of inequalities caused by multiple intersecting systems of marginalisation[10]The multilevel approach to modelling health inequalities at the intersection of multiple social dimensions provides a better understanding of the health heterogeneity existing in the population, and has been proposed as the gold standard for investigating health disparities in social epidemiology.[11]Despite this, its application to child health outcomes in sub-Saharan Africa remains limited.

This study uses nationally representative data from the 2024 Nigeria Demographic and Health Survey (NDHS) to examine the individual and structural determinants of ARI among children under five in Nigeria, with two primary aims: first, to estimate the associations between household cooking fuel type, socioeconomic factors, and child ARI using survey-weighted regression; and second, to apply MAIHDA to quantify the extent to which intersectional combinations of cooking fuel type, wealth, residence, and geopolitical zone produce clustered ARI risk beyond what individual-level factors alone can explain.

We hypothesise that: (1) polluting fuel use is independently associated with increased ARI risk, but that this association is substantially mediated by socioeconomic and geographic context; and (2) intersectional disadvantage — the co-occurrence of solid fuel use, poverty, rural residence, and geographic marginalisation — produces amplified ARI risk beyond the contribution of any single social position.

**Figure 1:**
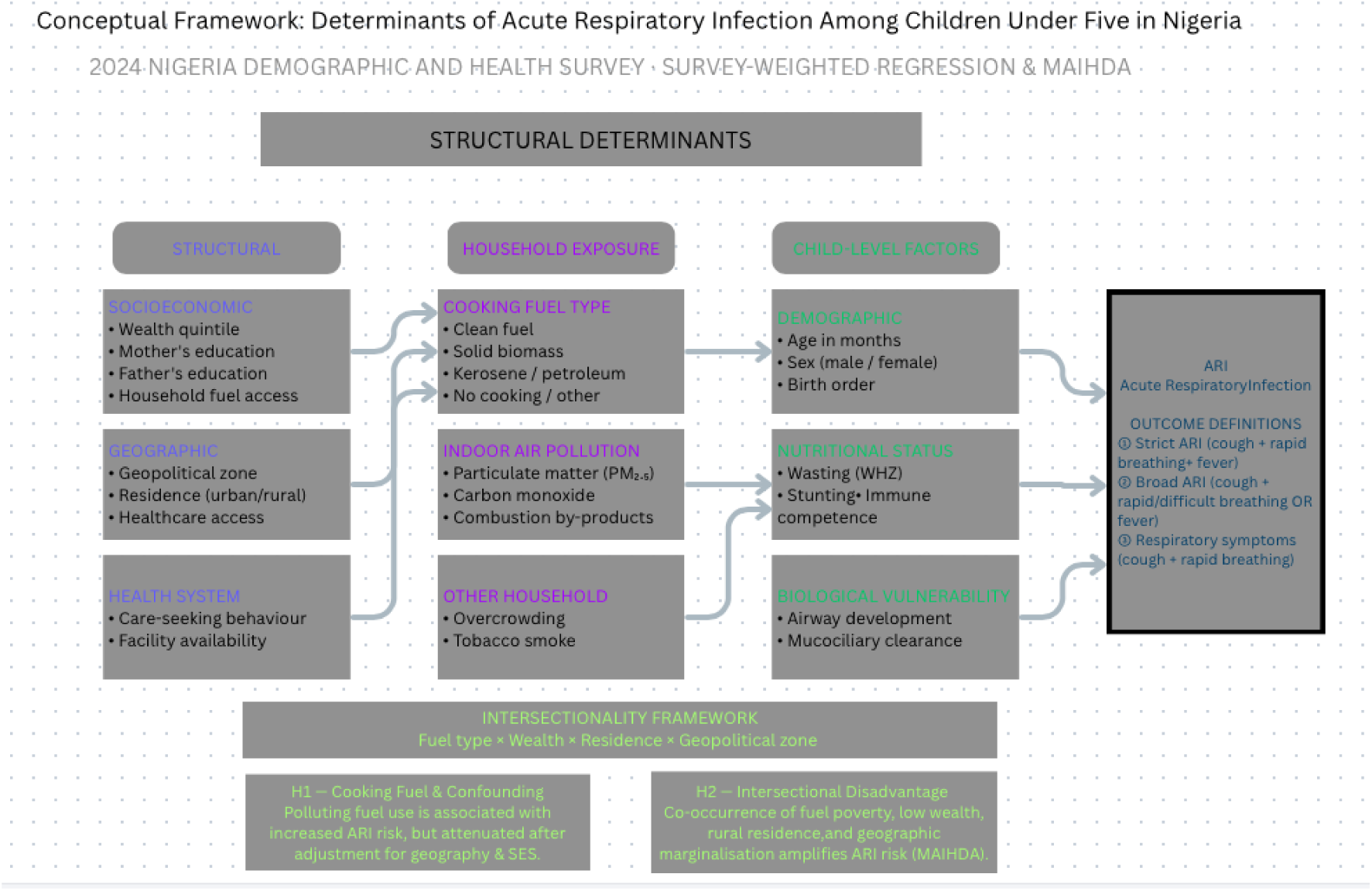
Conceptual Framework

## METHODS

### Study Design

This study is a secondary analysis of the 2024 Nigeria Demographic and Health Survey (NDHS), a nationally representative cross-sectional household survey. The NDHS employed a stratified two-stage cluster sampling design. Seventy-four sampling strata were created by stratifying each of Nigeria’s 36 states and the Federal Capital Territory into urban and rural areas.

In the first stage, enumeration areas (EAs) were selected within each stratum using probability proportional to size. In the second stage, households were systematically sampled within each selected EA following a household listing exercise. All analyses accounted for the complex survey design using sampling weights, clustering, and stratification variables provided in the dataset.

### Study Population

The analysis included children aged 0–59 months residing in interviewed households with available data on respiratory symptoms and household cooking fuel. We used the Children’s Recode (KR) file, which contains one record for each living child under five residing with the interviewed mother. Children were excluded if information was missing for the primary outcome (acute respiratory infection symptoms) or key exposure and confounding variables, including cooking fuel type, household wealth, geopolitical zone, or place of residence.

### Study Variables

#### Outcome: Acute Respiratory Infection (ARI)

The primary outcome was symptoms consistent with acute respiratory infection (ARI) within the two weeks preceding the survey. ARI was defined as maternal report of all three of the following symptoms: cough, rapid or difficult breathing, and fever. This definition aligns with the Integrated Management of Childhood Illness framework and prioritizes specificity for acute infectious respiratory illness. Children whose caregivers reported all three symptoms were classified as having ARI. To assess the robustness of findings to outcome definition, two alternative definitions were examined. The first classified children as having ARI if they had cough and either rapid breathing or fever, capturing possible cases with incomplete symptom reporting. The second definition included cough with rapid breathing but no fever, which may reflect non-febrile respiratory conditions. The strict definition (cough, rapid breathing, and fever) was used in all primary analyses.

#### Primary Exposure: Household Cooking Fuel

The primary exposure was the household’s main cooking fuel. Fuel types were grouped according to expected exposure to household air pollution. Categories included clean fuels (electricity, solar energy, liquefied petroleum gas, natural gas, and biogas), solid biomass fuels (wood, charcoal, sawdust, animal dung, agricultural crop residues, and biomass pellets), kerosene or other petroleum products (including kerosene, paraffin, and diesel), and other or no cooking in the household. Clean fuels served as the reference category in all analyses.

#### Covariates

Covariates were selected a priori based on established associations with childhood respiratory illness and household energy use. Child-level characteristics included age in months, categorized as <12 months, 12–23 months, and 24–59 months, and sex (male or female).

Household-level characteristics included wealth quintile (poorest, poorer, middle, richer, richest), derived from the DHS asset index; residence (urban or rural); and geopolitical zone (North Central, North East, North West, South East, South South, and South West).

Parental education included mother’s education (no education, primary, secondary, or higher) and father’s education (no education, primary, secondary, higher, or don’t know).

Nutritional status was assessed using wasting, defined as weight-for-height Z-score less than −2 standard deviations according to WHO growth standards. Anthropometric measurements were available for a subsample of children present during assessment (n = 9,402; 33%). Wasting was examined in secondary models restricted to this subsample.

#### Statistical Analysis

All analyses were conducted in R (version 4.4.1).

#### Design-Based Analysis

Primary analyses followed a design-based inference approach to preserve national representativeness. Survey-weighted analyses were conducted using the survey package. Sampling weights were normalized by dividing variable v005 by 1,000,000. Primary sampling units and survey strata were incorporated to account for clustering and stratification.

Weighted prevalence estimates and 95% confidence intervals were calculated for ARI across exposure categories and covariates. Associations between cooking fuel type and ARI were assessed using survey-weighted logistic regression models with a quasibinomial specification to accommodate potential overdispersion. Both unadjusted models and multivariable models adjusting for child age, sex, household wealth, residence, geopolitical zone, and mother’s education are presented. Results are reported as odds ratios with 95% confidence intervals and Wald p-values. Wasting was introduced in a separate model restricted to children with anthropometric measurements to examine its association with ARI without altering the primary analytic sample. Design-based inference was prioritized to ensure that estimates reflect the intended target population of Nigerian children under five years of age. Population-attributable fractions (PAF) and prevented fractions were calculated using the formula PAF = Pe(OR−1) / [1 + Pe(OR−1)], where Pe is the observed proportion exposed in the analytic sample and OR is the adjusted odds ratio. For protective exposures (OR < 1) this formula yields a negative value interpreted as the prevented fraction.

#### Multilevel Analysis of Individual Heterogeneity and Discriminatory Accuracy (MAIHDA)

Initial attempts to model interactions between cooking fuel and socioeconomic or geographic factors using conventional logistic regression with interaction terms resulted in unstable or inestimable coefficients due to sparse data and the low prevalence of ARI across many combinations of exposure and covariates. To address this, we conducted a complementary multilevel analysis using the MAIHDA framework. Intersectional strata were defined by combinations of cooking fuel type (four categories), wealth quintile (five categories), residence (two categories), and geopolitical zone (six categories), yielding 164 empirically observed strata out of 240 possible combinations. Many strata contained few children, and a substantial proportion had no ARI cases, reflecting the uneven distribution of exposure and risk across social and geographic groups. Two generalized linear mixed models with random intercepts for intersectional strata were estimated. The first model quantified variation in ARI across strata. The second additionally adjusted for child age and sex to distinguish basic demographic differences from variation associated with combinations of household-level social positions. Other covariates used in the regression models were not included as fixed effects in the MAIHDA models because they define the intersectional strata themselves.

Between-strata variance, the proportion of total variation attributable to intersectional group membership, and changes in variance after adjustment were examined to assess the extent to which ARI risk clustered across social configurations. Stratum-specific effects were derived from the estimated random effects to identify combinations associated with comparatively higher or lower risk.

#### Ethical Considerations

The NDHS protocol received ethical approval from the National Health Research Ethics Committee of Nigeria and the ICF Institutional Review Board. Informed consent was obtained from all respondents. This secondary analysis used de-identified, publicly available data and did not require additional ethical review.

## Results

**Table 1:**
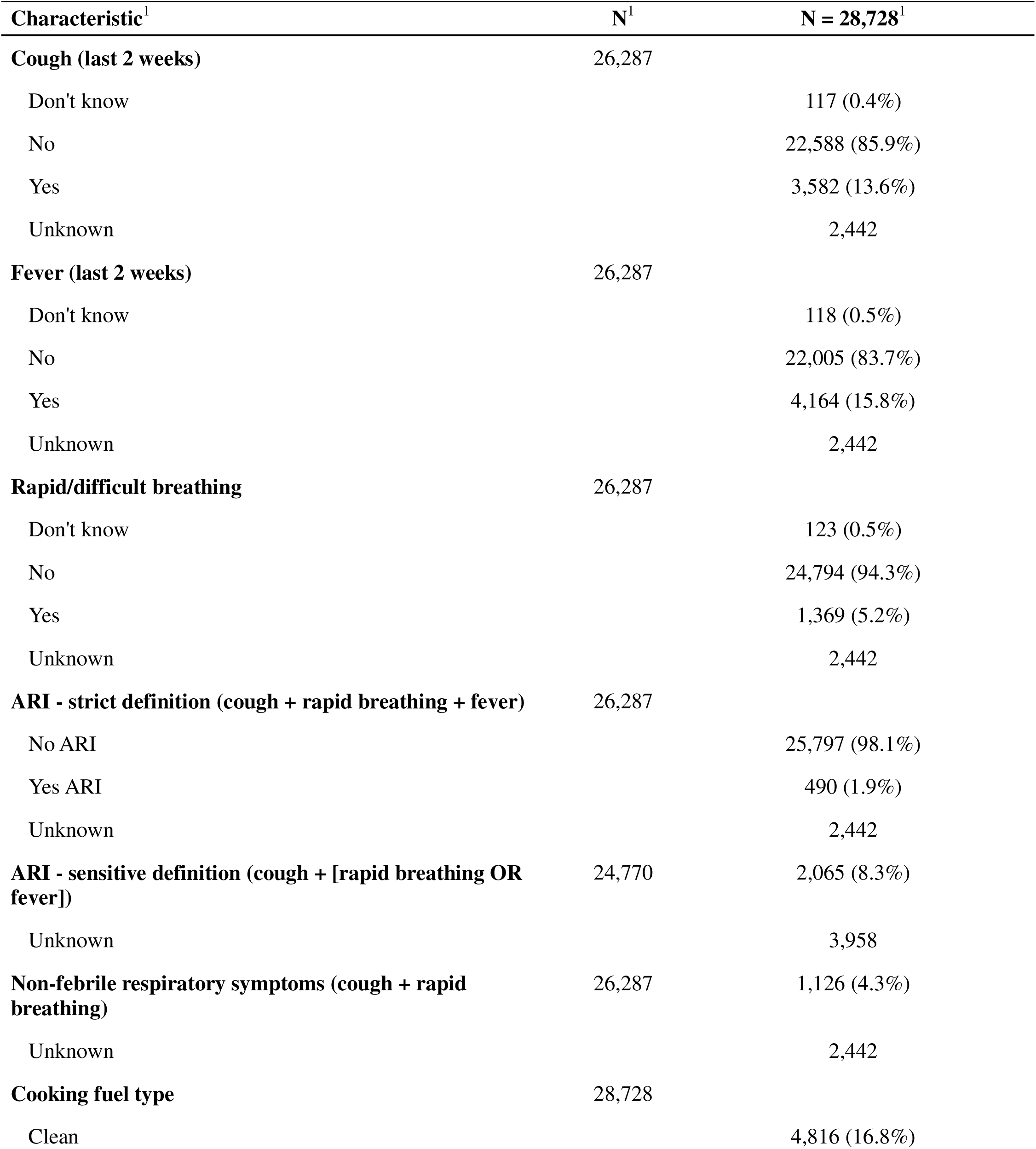
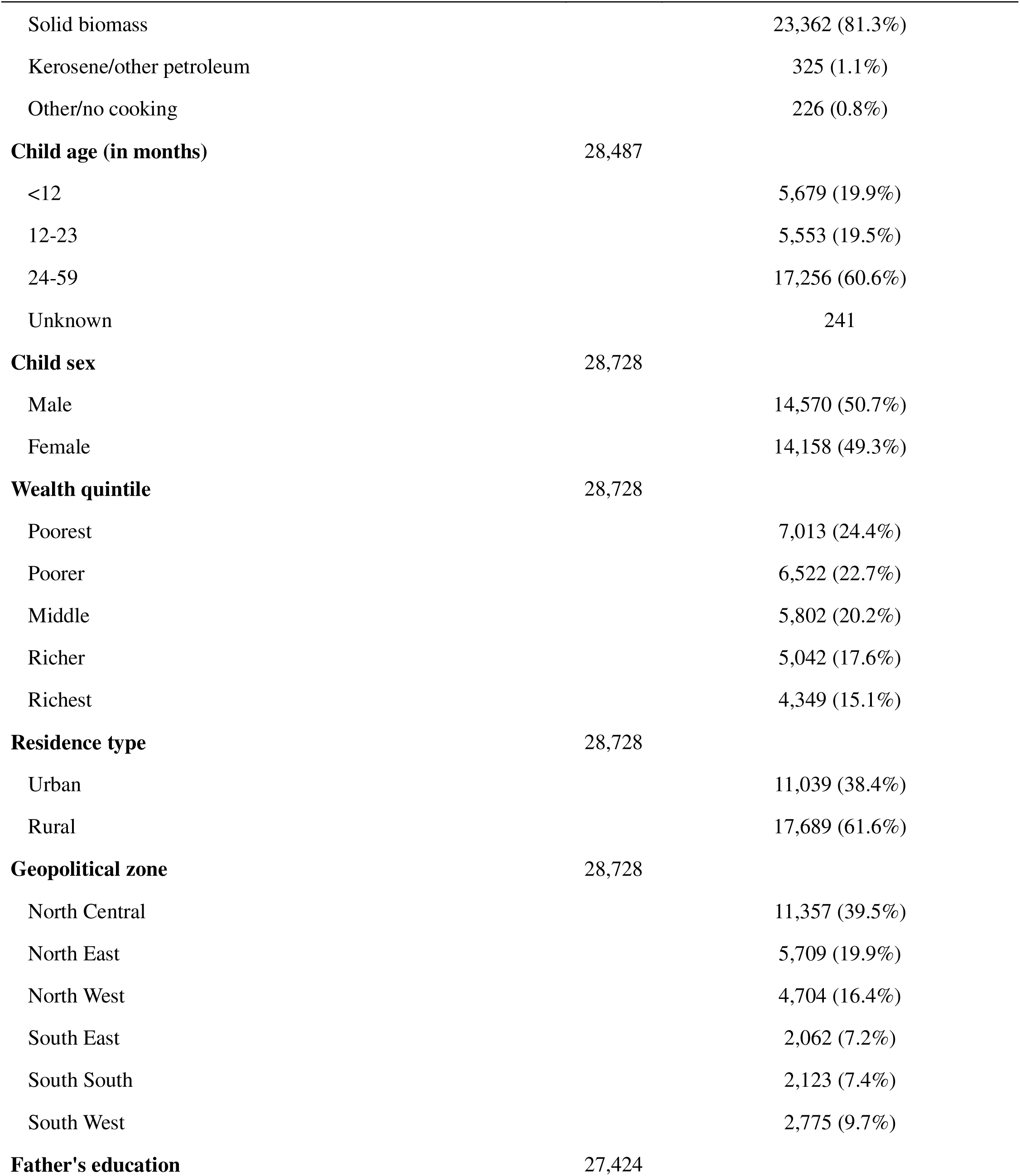
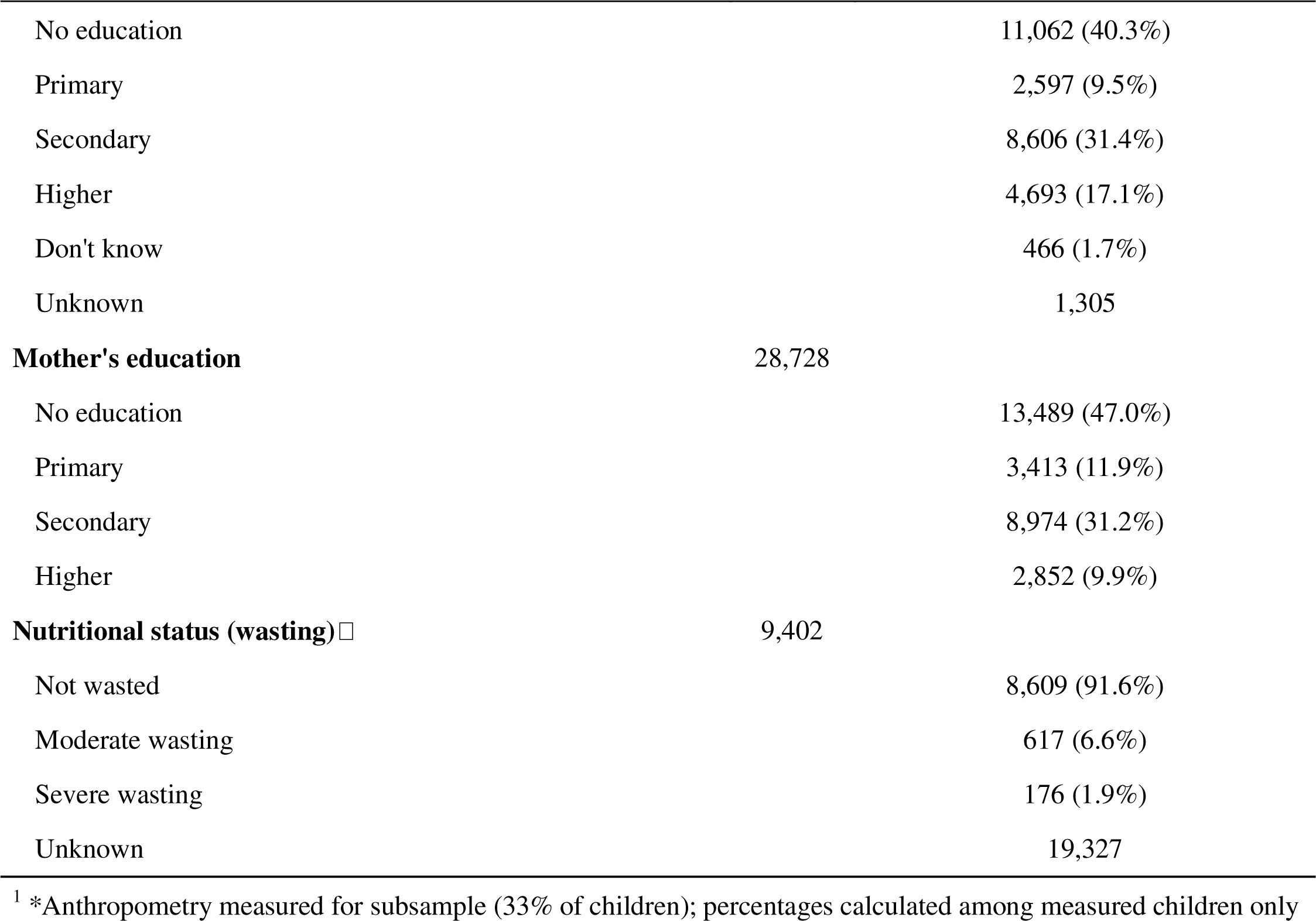
Sociodemographic characteristics and summary statistics of respondents.

| Characteristic<sup>1</sup> | N<sup>1</sup> | N = 28,728<sup>1</sup> |
| --- | --- | --- |
| **Cough (last 2 weeks)** | 26,287 |  |
| Don't know |  | 117 (0.4%) |
| No |  | 22,588 (85.9%) |
| Yes |  | 3,582 (13.6%) |
| Unknown |  | 2,442 |
| **Fever (last 2 weeks)** | 26,287 |  |
| Don't know |  | 118 (0.5%) |
| No |  | 22,005 (83.7%) |
| Yes |  | 4,164 (15.8%) |
| Unknown |  | 2,442 |
| **Rapid/difficult breathing** | 26,287 |  |
| Don't know |  | 123 (0.5%) |
| No |  | 24,794 (94.3%) |
| Yes |  | 1,369 (5.2%) |
| Unknown |  | 2,442 |
| **ARI - strict definition (cough + rapid breathing + fever)** | 26,287 |  |
| No ARI |  | 25,797 (98.1%) |
| Yes ARI |  | 490 (1.9%) |
| Unknown |  | 2,442 |
| **ARI - sensitive definition (cough + [rapid breathing OR fever])** | 24,770 | 2,065 (8.3%) |
| Unknown |  | 3,958 |
| **Non-febrile respiratory symptoms (cough + rapid breathing)** | 26,287 | 1,126 (4.3%) |
| Unknown |  | 2,442 |
| **Cooking fuel type** | 28,728 |  |
| Clean |  | 4,816 (16.8%) |

| **Characteristic<sup>1</sup>** | **N<sup>1</sup>** | **N = 28,728<sup>1</sup>** |
| --- | --- | --- |
| Solid biomass |  | 23,362 (81.3%) |
| Kerosene/other petroleum |  | 325 (1.1%) |
| Other/no cooking |  | 226 (0.8%) |
| **Child age (in months)** | **28,487** |  |
| <12 |  | 5,679 (19.9%) |
| 12-23 |  | 5,553 (19.5%) |
| 24-59 |  | 17,256 (60.6%) |
| Unknown |  | 241 |
| **Child sex** | **28,728** |  |
| Male |  | 14,570 (50.7%) |
| Female |  | 14,158 (49.3%) |
| **Wealth quintile** | **28,728** |  |
| Poorest |  | 7,013 (24.4%) |
| Poorer |  | 6,522 (22.7%) |
| Middle |  | 5,802 (20.2%) |
| Richer |  | 5,042 (17.6%) |
| Richest |  | 4,349 (15.1%) |
| **Residence type** | **28,728** |  |
| Urban |  | 11,039 (38.4%) |
| Rural |  | 17,689 (61.6%) |
| **Geopolitical zone** | **28,728** |  |
| North Central |  | 11,357 (39.5%) |
| North East |  | 5,709 (19.9%) |
| North West |  | 4,704 (16.4%) |
| South East |  | 2,062 (7.2%) |
| South South |  | 2,123 (7.4%) |
| South West |  | 2,775 (9.7%) |
| **Father's education** | **27,424** |  |
| No education |  | 11,062 (40.3%) |
| Primary |  | 2,597 (9.5%) |
| Secondary |  | 8,606 (31.4%) |
| Higher |  | 4,693 (17.1%) |
| Don't know |  | 466 (1.7%) |
| Unknown |  | 1,305 |
| **Mother's education** | **28,728** |  |
| No education |  | 13,489 (47.0%) |
| Primary |  | 3,413 (11.9%) |
| Secondary |  | 8,974 (31.2%) |
| Higher |  | 2,852 (9.9%) |
| **Nutritional status (wasting)□** | **9,402** |  |
| Not wasted |  | 8,609 (91.6%) |
| Moderate wasting |  | 617 (6.6%) |
| Severe wasting |  | 176 (1.9%) |
| Unknown |  | 19,327 |

<sup>1</sup> \*Anthropometry measured for subsample (33% of children); percentages calculated among measured children only

The analytic sample comprised 28,728 children under five. The majority lived in rural areas (61.6%) and were aged 24–59 months (60.6%), with roughly equal sex distribution. Nearly half of mothers (47.0%) and fathers (40.3%) had no formal education. Most households used polluting solid biomass fuel (81.3%), with only 16.8% using clean fuel. Wealth was skewed toward lower quintiles, with the poorest and poorer quintiles together accounting for 47.1% of the sample. The North Central zone was most represented (39.5%), reflecting the survey design. Anthropometric data were available for 9,402 children (32.7%); among these, 8.4% were wasted (6.5% moderate, 1.9% severe).

Regarding respiratory outcomes, 13.6% of children had cough in the preceding two weeks. Strict ARI (cough, rapid breathing, and fever) was present in 1.9% of children, while the broader respiratory symptom definition (cough with rapid/difficult breathing or fever) yielded a prevalence of 8.3%. Respiratory symptoms without fever occurred in 4.3%

**Table 2:** Unadjusted associations with ARI symptoms (survey-weighted quasibinomial logistic regression)

| Characteristic | ARI |  |  |  | Sensitivity<br>(Respiratory symptoms) |  |  |  |
| --- | --- | --- | --- | --- | --- | --- | --- | --- |
|  | N | OR<sup>1</sup> | 95% CI<sup>1</sup> | p-value | N | OR<sup>1</sup> | 95% CI<sup>1</sup> | p-value |
| **Child (In months)** | 26,064 |  |  | **<0.001** | 26,064 |  |  | **<0.001** |
| < 12 |  | — | — |  |  | — | — |  |
| 12-23 |  | 0.87 | 0.63, 1.20 | 0.402 |  | 0.86 | 0.70, 1.06 | 0.161 |
| 24-59 |  | 0.52 | 0.40, 0.68 | **<0.001** |  | 0.61 | 0.51, 0.72 | **<0.001** |
| **Cooking fuel type** | 26,286 |  |  | 0.244 | 26,286 |  |  | **<0.001** |
| Clean |  | — | — |  |  | — | — |  |
| Polluting fuel |  | 1.40 | 0.98, 2.01 | 0.068 |  | 1.85 | 1.43, 2.39 | **<0.001** |
| Kerosene/other petroleum |  | 1.14 | 0.49, 2.64 | 0.766 |  | 0.97 | 0.45, 2.10 | 0.932 |
| No cooking/Other |  | 0.81 | 0.25, 2.67 | 0.733 |  | 1.56 | 0.70, 3.48 | 0.281 |
| **Child sex** | 26,286 |  |  | 0.945 | 26,286 |  |  | 0.815 |
| Male |  | — | — |  |  | — | — |  |
| Female |  | 1.01 | 0.80, 1.27 | 0.945 |  | 1.02 | 0.88, 1.18 | 0.815 |
| **Wealth quintile** | 26,286 |  |  | 0.567 | 26,286 |  |  | **0.004** |
| Poorest |  | — | — |  |  | — | — |  |
| Poorer |  | 1.11 | 0.81, 1.51 | 0.522 |  | 0.95 | 0.74, 1.22 | 0.698 |

| Characteristic | N | ARI |  |  | N | Sensitivity<br>(Respiratory symptoms) |  |  |
| --- | --- | --- | --- | --- | --- | --- | --- | --- |
|  |  | OR<sup>1</sup> | 95% CI<sup>1</sup> | p-value |  | OR<sup>1</sup> | 95% CI<sup>1</sup> | p-value |
| Middle |  | 1.12 | 0.81, 1.55 | 0.476 |  | 0.85 | 0.66, 1.09 | 0.193 |
| Richer |  | 0.83 | 0.57, 1.21 | 0.332 |  | 0.72 | 0.54, 0.96 | **0.027** |
| Richest |  | 0.93 | 0.62, 1.38 | 0.705 |  | 0.56 | 0.42, 0.76 | **<0.001** |
| **Residence type** | 26,286 |  |  | 0.581 | 26,286 |  |  | 0.676 |
| Urban |  | — | — |  |  | — | — |  |
| Rural |  | 1.07 | 0.84, 1.37 | 0.581 |  | 0.96 | 0.78, 1.17 | 0.676 |
| **Geopolitical zone** | 26,286 |  |  | **<0.001** | 26,286 |  |  | **<0.001** |
| North Central |  | — | — |  |  | — | — |  |
| North East |  | 1.08 | 0.81, 1.46 | 0.594 |  | 1.44 | 1.12, 1.84 | **0.004** |
| North West |  | 0.46 | 0.32, 0.66 | **<0.001** |  | 0.52 | 0.40, 0.69 | **<0.001** |
| South East |  | 0.63 | 0.41, 0.97 | **0.034** |  | 0.51 | 0.36, 0.74 | **<0.001** |
| South South |  | 0.95 | 0.67, 1.34 | 0.762 |  | 0.63 | 0.47, 0.86 | **0.004** |
| South West |  | 0.24 | 0.14, 0.43 | **<0.001** |  | 0.22 | 0.14, 0.33 | **<0.001** |
| **Father's education level** | 25,084 |  |  | 0.570 | 25,084 |  |  | 0.789 |
| No education |  | — | — |  |  | — | — |  |
| Primary |  | 1.03 | 0.68, 1.56 | 0.907 |  | 0.91 | 0.68, 1.23 | 0.539 |
|  |  | OR<sup>1</sup> | 95% CI<sup>1</sup> | p-value |  | OR<sup>1</sup> | 95% CI<sup>1</sup> | p-value |
| Secondary |  | 1.24 | 0.92, 1.69 | 0.158 |  | 0.93 | 0.74, 1.16 | 0.521 |
| Higher |  | 1.07 | 0.76, 1.51 | 0.696 |  | 0.86 | 0.67, 1.10 | 0.220 |
| Don't know |  | 0.75 | 0.22, 2.58 | 0.650 |  | 0.83 | 0.40, 1.72 | 0.611 |
| **Mother's education** | 26,286 |  |  | 0.579 | 26,286 |  |  | **0.011** |
| No education |  | — | — |  |  | — | — |  |
| Primary |  | 0.95 | 0.66, 1.37 | 0.788 |  | 0.79 | 0.60, 1.05 | 0.101 |
| Secondary |  | 1.05 | 0.80, 1.38 | 0.722 |  | 0.88 | 0.71, 1.09 | 0.248 |
| Higher |  | 0.80 | 0.53, 1.19 | 0.275 |  | 0.63 | 0.48, 0.84 | **0.002** |

<sup>1</sup>OR = Odds Ratio, CI = Confidence Interval

Child age was the most consistent predictor across both outcomes. Compared to infants under 12 months, children aged 24–59 months had significantly lower odds of both strict ARI (OR 0.52, 95% CI 0.40–0.68) and broader respiratory symptoms (OR 0.61, 95% CI 0.51–0.72). Cooking fuel showed a divergent pattern by outcome definition. Polluting fuel use was not significantly associated with strict ARI (OR 1.40, p=0.068), but was strongly associated with broader respiratory symptoms (OR 1.85, 95% CI 1.43–2.39, p<0.001). Geopolitical zone was significantly associated with both outcomes. Relative to North Central, children in North West and South West had markedly lower odds of both ARI (OR 0.46 and 0.24 respectively) and respiratory symptoms (OR 0.52 and 0.22). North East children had elevated odds of respiratory symptoms (OR 1.44, p=0.004) but not strict ARI. Wealth was significant only for respiratory symptoms (p=0.004), with Richer and Richest quintile children showing progressively lower odds compared to the poorest. Mother’s higher education was associated with reduced respiratory symptoms (OR 0.63, p=0.002) but not strict ARI.

**Table 3:** Adjusted associations with ARI symptoms (survey-weighted quasibinomial logistic regression)

| Characteristic | ARI<br>Adjusted |  |  | Respiratory symptoms<br>Adjusted |  |  |
| --- | --- | --- | --- | --- | --- | --- |
|  | OR<sup>1</sup> | 95% CI<sup>1</sup> | p-value | OR<sup>1</sup> | 95% CI<sup>1</sup> | p-value |
| **Cooking fuel type** |  |  | 0.6 |  |  | 0.8 |
| Clean (ref) | — | — |  | — | — |  |
| Polluting fuel | 1.35 | 0.77, 2.36 | 0.3 | 1.17 | 0.78, 1.76 | 0.4 |
| Kerosene/other petroleum | 1.15 | 0.48, 2.78 | 0.8 | 0.94 | 0.41, 2.18 | 0.9 |
| No cooking/Other | 0.77 | 0.22, 2.68 | 0.7 | 1.12 | 0.44, 2.86 | 0.8 |
| **Child age (in months)** |  |  | <0.001 |  |  | <0.001 |
| <12 (ref) | — | — |  | — | — |  |
| 12-23 | 0.88 | 0.64, 1.22 | 0.4 | 0.88 | 0.72, 1.08 | 0.2 |
| 24-59 | 0.53 | 0.41, 0.68 | <0.001 | 0.62 | 0.52, 0.74 | <0.001 |
| **Child sex** |  |  | >0.9 |  |  | 0.8 |
| Male (ref) | — | — |  | — | — |  |
| Female | 1.01 | 0.80, 1.28 | >0.9 | 1.02 | 0.88, 1.19 | 0.8 |
| **Wealth quintile** |  |  | 0.2 |  |  | 0.5 |
| Poorest (ref) | — | — |  | — | — |  |
| Poorer | 1.20 | 0.88, 1.64 | 0.3 | 1.01 | 0.78, 1.31 | >0.9 |
| Middle | 1.30 | 0.93, 1.81 | 0.13 | 0.87 | 0.67, 1.13 | 0.3 |
| Richer | 1.13 | 0.75, 1.71 | 0.6 | 0.78 | 0.56, 1.08 | 0.13 |
| Richest | 1.86 | 1.04, 3.33 | 0.037 | 0.85 | 0.53, 1.39 | 0.5 |
| **Residence type** |  |  | 0.8 |  |  | 0.003 |
| Urban (ref) | — | — |  | — | — |  |
| Rural | 0.96 | 0.73, 1.26 | 0.8 | 0.71 | 0.57, 0.89 | 0.003 |
| **Geopolitical zone** |  |  | <0.001 |  |  | <0.001 |
| North Central (ref) | — | — |  | — | — |  |
| North East | 1.10 | 0.82, 1.47 | 0.5 | 1.34 | 1.04, 1.71 | 0.021 |
| North West | 0.45 | 0.31, 0.65 | <0.001 | 0.51 | 0.39, 0.67 | <0.001 |
|  | OR<sup>1</sup> | 95% CI<sup>1</sup> | p-value | OR<sup>1</sup> | 95% CI<sup>1</sup> | p-value |
| South East | 0.60 | 0.38, 0.95 | **0.030** | 0.47 | 0.32, 0.69 | **<0.001** |
| South South | 0.89 | 0.55, 1.43 | 0.6 | 0.56 | 0.39, 0.81 | **0.002** |
| South West | 0.22 | 0.11, 0.44 | **<0.001** | 0.19 | 0.12, 0.31 | **<0.001** |
| **Mother's education** |  |  |  |  |  | 0.070 |
| No education (ref) |  |  |  | — | — |  |
| Primary |  |  |  | 0.98 | 0.74, 1.31 | 0.9 |
| Secondary |  |  |  | 1.35 | 1.05, 1.73 | **0.020** |
| Higher |  |  |  | 1.15 | 0.81, 1.62 | 0.4 |

<sup>1</sup>OR = Odds Ratio, CI = Confidence Interval

After adjustment, child age remained the only consistent individual-level predictor across both outcomes. Children aged 24–59 months had significantly lower odds of strict ARI (aOR 0.53, 95% CI 0.41–0.68) and respiratory symptoms (aOR 0.62, 95% CI 0.52–0.74) compared to infants under 12 months. Cooking fuel lost statistical significance in both adjusted models, with polluting fuel showing attenuated and non-significant associations for both strict ARI (aOR 1.35, p=0.3) and respiratory symptoms (aOR 1.17, p=0.4). North West and South West consistently showed the lowest odds relative to North Central, while in the respiratory symptoms model, North East had elevated odds (aOR 1.34, p=0.021) and all southern zones showed significantly reduced odds. Rural residence was significantly associated with lower odds of respiratory symptoms after adjustment (aOR 0.71, 95% CI 0.57–0.89, p=0.003) but not strict ARI, a reversal from the unadjusted null finding that warrants cautious interpretation.

An unexpected finding was that Richest wealth quintile children had higher adjusted odds of strict ARI compared to the poorest (aOR 1.86, p=0.037), despite no such association in the unadjusted model — suggesting possible negative confounding. Mother’s secondary education was associated with higher odds of respiratory symptoms (aOR 1.35, p=0.020), though the overall education term was only marginally significant (p=0.070).

To quantify the population-level relevance of key associations, population-attributable fractions (PAF) and prevented fractions (PF) were estimated using adjusted odds ratios and observed exposure prevalences. In unadjusted analysis, 41.4% of broad respiratory symptom cases were attributable to polluting fuel use; after adjustment for geographic and socioeconomic factors this fell to 12.4%, quantifying the degree to which the crude fuel-ARI association reflects structural confounding. Child age 24–59 months was the largest preventable factor: if all children carried the vulnerability of infants under 12 months, strict ARI burden would be an estimated 39.8% higher (broad symptoms: 29.9%). Rural residence was associated with a prevented fraction of 21.7% for broad respiratory symptoms after adjustment.

**Table 4:**
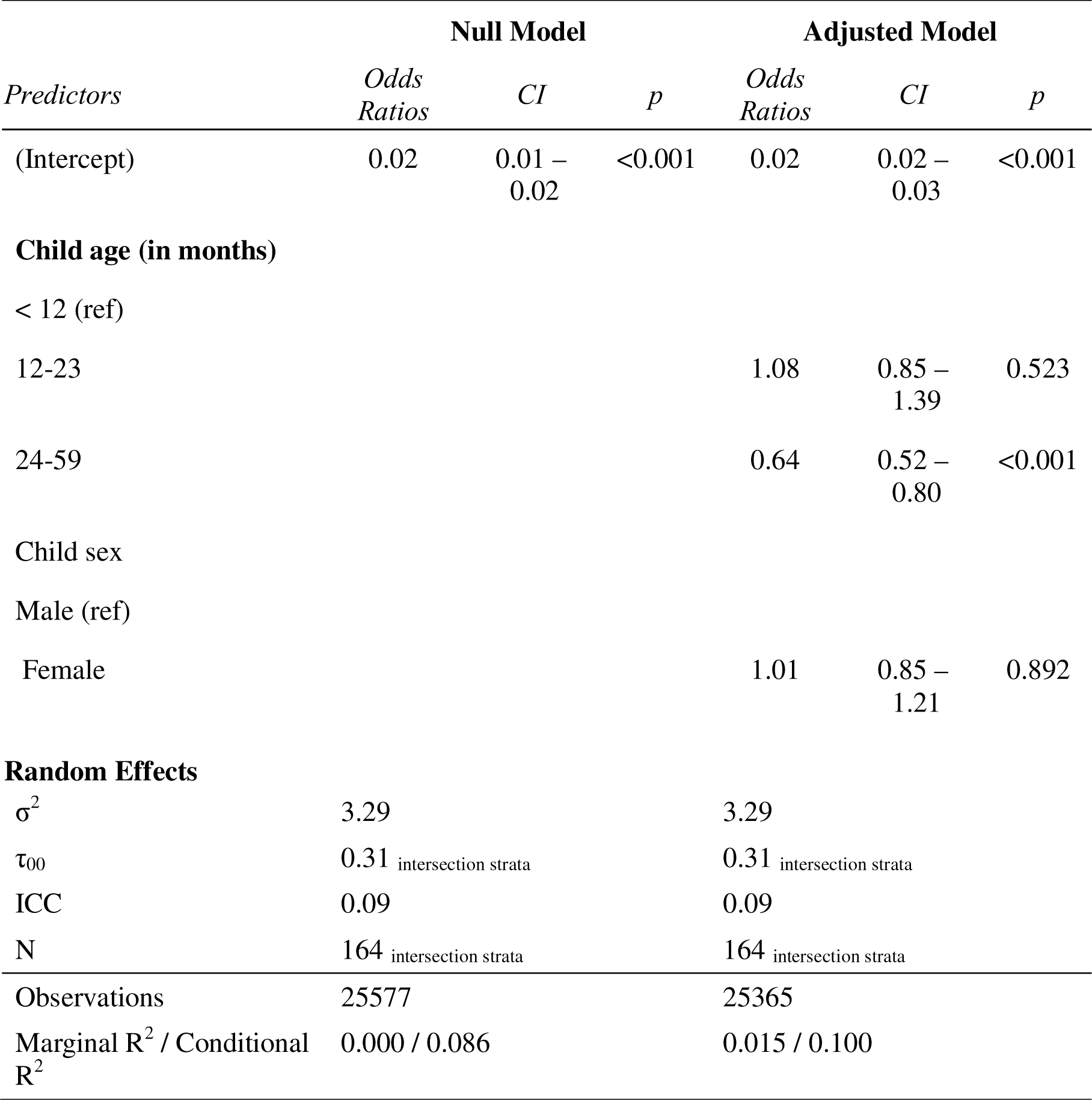
MAIHDA Results: ARI in Nigerian Children.

Adding child age and sex in the adjusted model explained a small proportion of between-strata variance (marginal R² increased from 0.000 to 0.015), while the conditional R² rose from 0.086 to 0.100, suggesting that intersectional strata account for substantially more variance than individual demographic characteristics alone. The between-strata variance was unchanged (τ = 0.31), indicating that age and sex do not explain the clustering of ARI risk across social strata. Older children (24–59 months) had lower odds of ARI (OR 0.64, 95% CI 0.52–0.80) consistent with the regression findings, while sex was not associated with ARI risk (OR 1.01, p=0.892)

**Figure 2:**
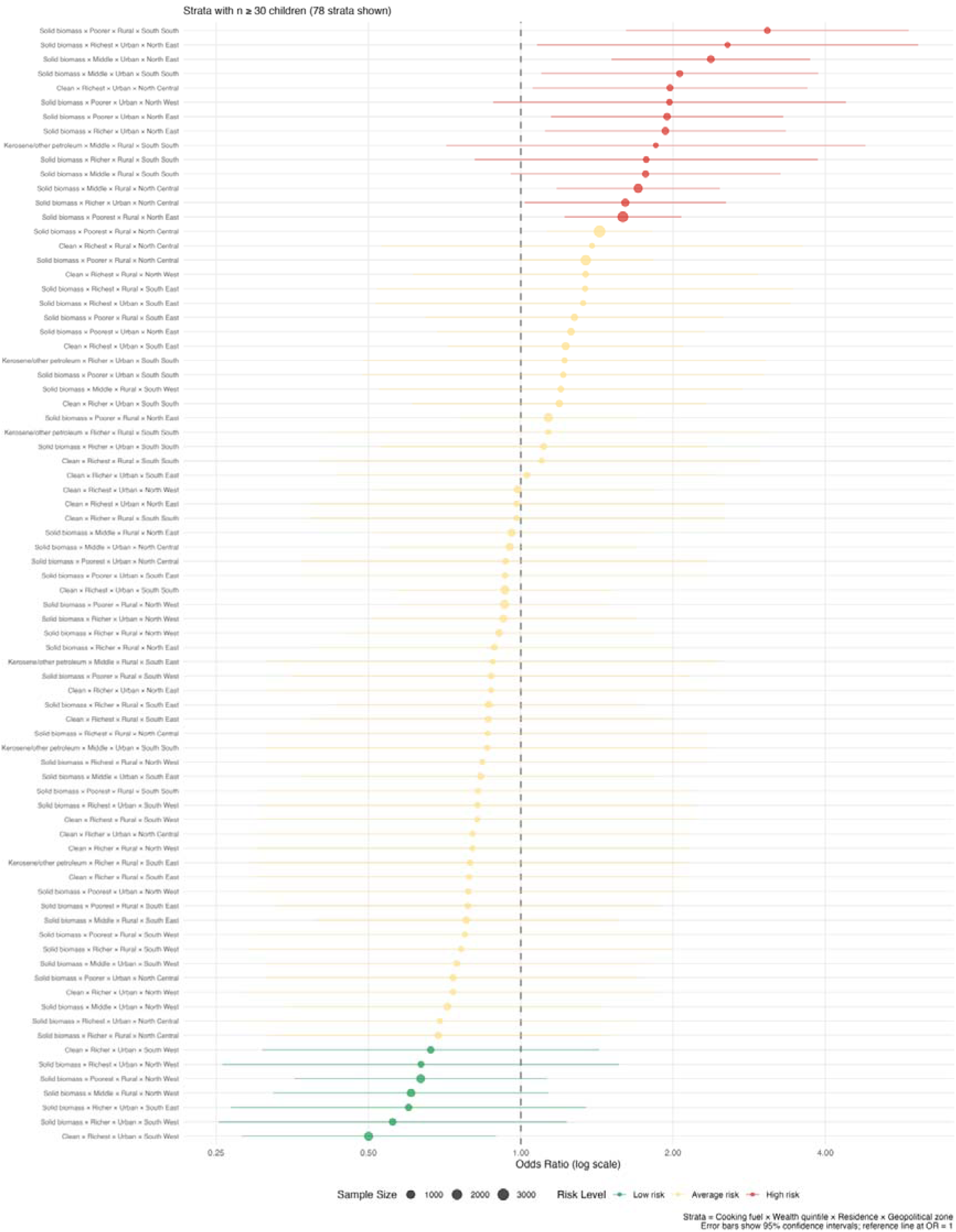
A caterpillar plot showing intersectional strata and ARI risk

This plot details the risk profiles for 76 specific intersectional strata with sample sizes under 30 children.

**Figure 3:**
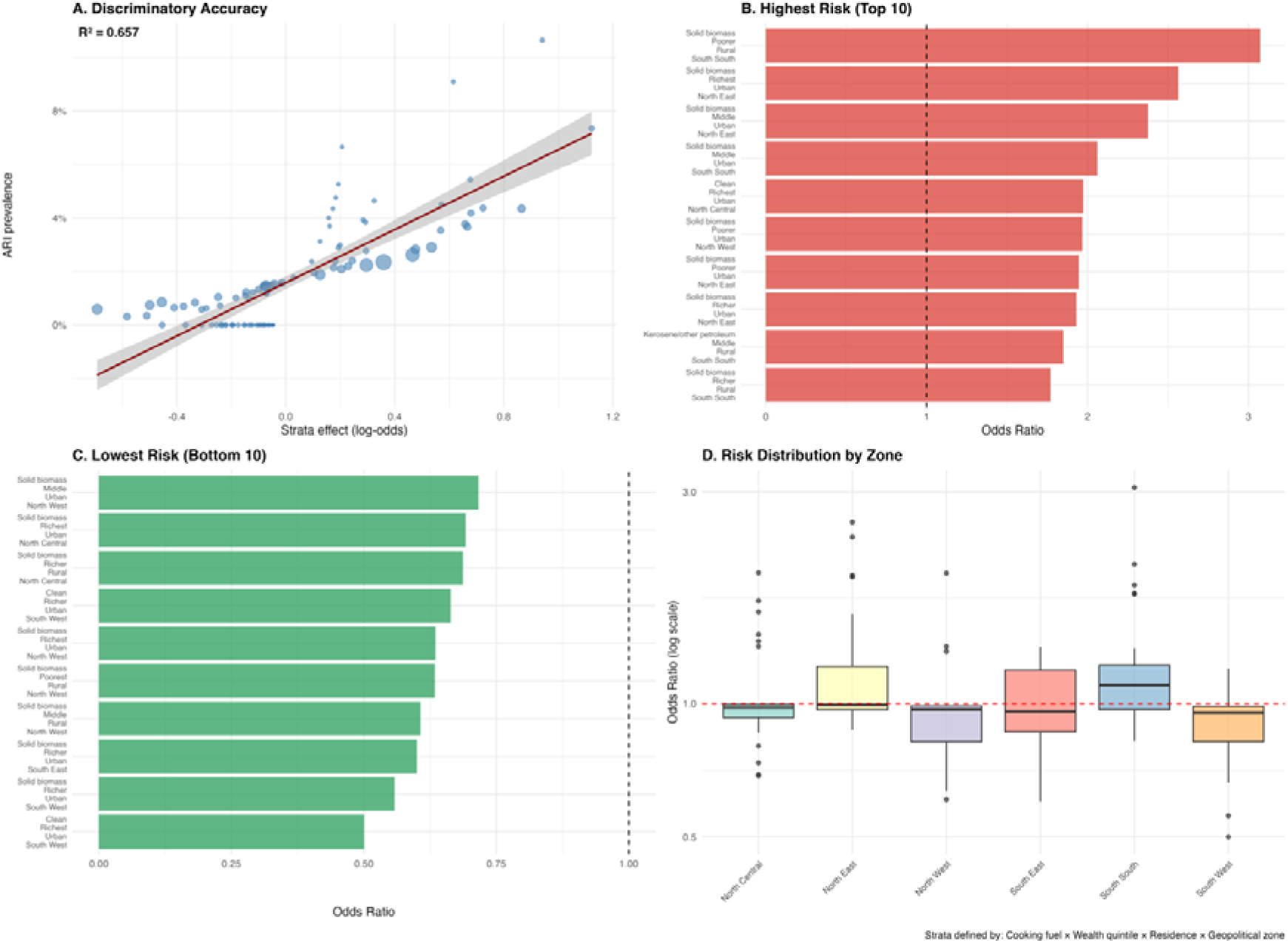
MAIHDA summary of intersectional Analysis of ARI in Nigerian Children

## Discussion

This study examined the determinants of acute respiratory infection among children under five in Nigeria, using nationally representative DHS data and a dual analytic strategy combining survey-weighted regression with MAIHDA. The findings contribute to growing evidence on the structural drivers of childhood respiratory illness in sub-Saharan Africa, with particular relevance for equity-focused policy.

### Household Air Pollution and Cooking Fuel

The association between solid biomass fuel use and childhood ARI has been widely documented across sub-Saharan Africa. A multilevel logistic analysis using Tanzania DHS data found that children in households using unclean fuels were substantially more likely to suffer from ARI than those using clean fuels.[12] Our findings partially support this relationship. In unadjusted analyses, polluting fuel use was strongly associated with broader respiratory symptoms (OR 1.85), consistent with biological plausibility: household air pollution has been demonstrated to impair phagocytosis by macrophages and adherence to surface, reduce the clearance of bacterial from the respiratory tract [i.e. mucociliary], and disrupt the alveolar-capillary barrier in the lungs.[13] However, this association was substantially attenuated and became non-significant after adjustment for geopolitical zone and socioeconomic factors. This pattern mirrors findings from broader regional analyses — a pooled study of 31 sub-Saharan African countries using DHS data found that the association between cooking fuel and ARI symptoms was modified by residence type and broader socioeconomic context[2] — suggesting that the crude fuel-ARI relationship is substantially confounded by the structural determinants of fuel access.

The divergence between outcome definitions is notable: cooking fuel was not significantly associated with strict ARI (fever plus cough plus rapid breathing) in either unadjusted or adjusted models yet showed a strong unadjusted association with the broader respiratory symptom definition. This suggests that household air pollution may contribute more to non-febrile respiratory morbidity — cough and breathing difficulties without systemic infection — than to clinically confirmed febrile ARI. Respiratory infections comprising both upper and lower respiratory tract infections have all been associated with exposure to household air pollution though the severity gradient may differ by outcome definition.[14] This distinction has implications for how household air pollution interventions are evaluated; trials using strict febrile ARI endpoints may underestimate the respiratory burden attributable to biomass fuel exposure. The PAF for polluting fuel fell from 41.4% to 12.4% after adjustment, numerically illustrating how much of the apparent fuel burden is attributable to the structural conditions that determine fuel access rather than fuel exposure itself.

Younger children, particularly infants under 12 months, consistently had the highest odds of ARI across all models and both outcome definitions. Children aged 2–23 months have shown the highest prevalence of ARI in Nigerian hospital-based studies, particularly those aged 2–11 months who showed the highest prevalence of severe pneumonia, a pattern supported by evidence of increased vulnerability in infants due to immature immune systems and underdeveloped airways.[15] The protective effect of older age (24–59 months, aOR 0.53 for strict ARI) is consistent with this and has been observed across DHS-based analyses in the region. Studies across 37 sub-Saharan African countries have observed a consistent correlation between younger age and higher ARI risk, with immunity accumulating through exposure over the first years of life.[16] These findings reinforce the importance of targeting ARI prevention and case management efforts toward the youngest children, particularly in the first year of life.

Geopolitical zone was the most robust predictor of ARI in both unadjusted and adjusted models, with North West and South West zones showing markedly lower odds relative to North Central across both outcome definitions. These zone-level effects persisted after controlling for individual and household characteristics, pointing to unmeasured contextual factors operating at the subnational level. Spatial analysis of ARI in Nigeria using 2013 DHS data identified significant hotspots in North East states, attributed to factors including prevailing dust-laden north-east trade winds and healthcare system challenges in conflict-affected areas.[9] The consistently lower odds in North West and South West may be explained by differences in healthcare-seeking behaviour, climatic factors, and diagnostic practices rather than truly lower disease burden. Healthcare utilisation in Nigeria is consistently lower among those living in North West and North East geopolitical zones, suggesting that reporting bias due to reduced health care-seeking behaviour may explain lower observed ARI prevalence in these regions.[17] The prevented fraction of 39.8% for strict ARI associated with older age underscores that the youngest infants represent the highest-yield target group for ARI prevention efforts.

Three counterintuitive findings warrant discussion. First, children in the richest wealth quintile had higher adjusted odds of strict ARI compared to the poorest (aOR 1.86), despite no such association in unadjusted analyses. This likely reflects negative confounding by geopolitical zone: wealthier households are disproportionately concentrated in zones where strict ARI prevalence is lower, and adjusting for zone unmasks a within-zone wealth effect. Alternatively, wealthier households may have greater healthcare access, leading to higher rates of care-seeking and thus more captured ARI episodes in caregiver recall. Evidence from sub-Saharan Africa suggests that wealthier populations may paradoxically benefit more from health interventions due to differential access, which can complicate interpretation of wealth-ARI associations in cross-sectional data.[16,17]

Second, rural residence was associated with lower odds of broader respiratory symptoms after adjustment, despite a null unadjusted finding. This counterintuitive result may reflect differential healthcare-seeking and symptom recognition between urban and rural caregivers. The relationship between urbanicity and ARI risk is complex, as urban households experience different indoor pollution dynamics and overcrowding patterns compared to rural settings and urban mothers may report respiratory symptoms more readily due to greater health literacy and facility access.[2]

Third, maternal secondary education was associated with higher odds of respiratory symptoms in the adjusted model while higher education showed a non-significant protective trend. This paradox may reflect a surveillance effect — mothers with secondary education may be more likely to recognise and report respiratory symptoms without necessarily having higher true disease burden. It may also reflect residual confounding by unmeasured community-level characteristics correlated with secondary but not higher education attainment in Nigeria’s specific educational context.

The MAIHDA analysis revealed that approximately 9% of total variance in ARI occurrence was attributable to differences between intersectional strata defined by cooking fuel, wealth, residence, and geopolitical zone. Intersectional MAIHDA is an innovative approach for investigating inequalities in health outcomes, with conceptual and methodological advantages over conventional single-level regression analysis, including the ability to handle sparse strata through partial pooling and to quantify between-strata variance without pre-specifying interaction terms.[18] While an ICC of 0.09 is modest in absolute terms, it is substantively meaningful in the context of a low-prevalence binary outcome, and is consistent with comparable applications. A MAIHDA study of under-five health outcomes in Nairobi slums found comparable discriminatory accuracy for diarrhoea (VPC = 9.0%), with the authors concluding that intersectional strata meaningfully differentiate children at higher risk.[19] Critically, adding child age and sex to the adjusted model did not reduce between-strata variance (τ unchanged at 0.31), confirming that the clustering of ARI risk across social strata is not explained by the demographic composition of those strata. This finding supports an intersectional interpretation: the combination of fuel type, wealth, residence, and geography produces patterned health disadvantage that operates independently of individual child characteristics. The ICC in MAIHDA is a measure of discriminatory accuracy, indicating the share of total individual variance at the stratum level — the higher the ICC, the better the model discriminates individuals with higher or lower outcome risk across intersectional strata.[20]

Anthropometric data were available for 32.7% of the sample (n = 9,401, see appendix), reflecting the DHS subsample measurement design. Among measured children, wasting prevalence was 8.4%, consistent with national estimates. In unadjusted analyses, wasting was not significantly associated with strict ARI (p = 0.159), though the direction of estimates — moderately wasted children appearing to have lower ARI risk than non-wasted children (OR 2.06, p = 0.063, reference: moderate wasting) — likely reflects limited statistical power given the rarity of strict ARI within an already restricted subsample, and possible survivor bias. Nigerian-specific analyses have highlighted malnutrition as a significant contributor to ARI risk, and future studies with complete anthropometric coverage would better capture this relationship.[21]

### Limitations

This study has several limitations. The cross-sectional design precludes causal inference. ARI was defined using caregiver-reported symptoms over a two-week recall period, introducing potential recall and social desirability bias. The divergent findings across strict and broad outcome definitions highlight sensitivity to case definition, and neither definition constitutes a clinical diagnosis. Survey weights were incorporated in regression analyses to ensure national representativeness, but MAIHDA models were unweighted, limiting direct comparability; the MAIHDA findings should therefore be treated as exploratory. Finally, unmeasured confounders including indoor ventilation quality, tobacco smoke exposure, vaccination status, and healthcare-seeking behaviour may explain residual associations - individual vaccination status was not included in the primary intersectional strata construction to maintain model parsimony and statistical power, the inclusion of maternal education and geopolitical zone serves as a robust proxy for the structural barriers to immunization access prevalent in Nigeria.

## Data Availability

The primary data are from the 2024 Nigeria Demographic and Health Survey, accessible via The DHS Program after free registration and project approval. The R code, scripts, processed data, and outputs are openly available at https://doi.org/10.5281/zenodo.18832992 version 1.0 (linked to https://github.com/olasuraheem/NDHS_ARI), under MIT license, with reproduction instructions in the README.

## List of Abbreviations

ARI: Acute Respiratory Infection
aOR: Adjusted Odds Ratio
CI: Confidence Interval
DHS: Demographic and Health Survey
HAP: Household Air Pollution
ICC: Intraclass Correlation Coefficient
LMIC: Low- and Middle-Income Country
MAIHDA: Multilevel Analysis of Individual Heterogeneity and Discriminatory Accuracy
NDHS: Nigeria Demographic and Health Survey
OR: Odds Ratio
PAF: Population-Attributable Fraction
PCV: Proportional Change in Variance
PF: Prevented Fraction
PM.: Particulate Matter ≤2.5 micrometres
SES: Socioeconomic Status
WHZ: Weight-for-Height Z-score
WHO: World Health Organization

## Declarations

### Ethics approval and consent to participate

This study is a secondary analysis of publicly available data from the 2024 Nigeria Demographic and Health Survey (2024 NDHS). Ethical approval for the NDHS was obtained from the ICF Institutional Review Board and the National Health Research Ethics Committee (NHREC) of Nigeria prior to data collection. All survey participants provided informed consent before participation. As this study involved analysis of fully anonymized, publicly available data with no direct contact with human participants, no additional institutional ethical approval was required. Data access was approved following registration with The DHS Program.

### Consent for publication

Not applicable.

### Authors’ contributions

KSA conceptualised the study, developed the methodology, conducted the formal analysis, performed data curation and visualisation, and wrote the original draft. MTO contributed to data interpretation and critical revision of the manuscript. BAM contributed to critical revision of the manuscript and review of the final draft. ATA contributed to the review and editing of the manuscript. ISA contributed to supervision and critical revision of the manuscript. All authors read and approved the final manuscript.

## Acknowledgements

The authors gratefully acknowledge the DHS Program for granting access to the 2024 Nigeria Demographic and Health Survey data. The views expressed in this article are those of the authors alone and do not represent the DHS Program or any affiliated organisation.

## Authors’ information

Kamaldeen Sunkanmi Abdulraheem is a physician at the Department of Community Health and Primary Care, Lagos University Teaching Hospital, Lagos, Nigeria. Correspondence.

